# SIGNIFICANT IMPROVEMENT IN MELANOMA SURVIVAL OVER THE LAST DECADE: A HUNGARIAN NATIONWIDE STUDY BETWEEN 2011–2019

**DOI:** 10.1101/2022.04.06.22273390

**Authors:** Gabriella Liszkay, Angela Benedek, Csaba Polgár, Judit Oláh, Péter Holló, Gabriella Emri, András Csejtei, István Kenessey, Zoltán Polányi, Kata Knollmajer, Máté Várnai, Zoltán Vokó, Balázs Nagy, György Rokszin, Ibolya Fábián, Zsófia Barcza, Rolland Gyulai, Zoltan Kiss

## Abstract

**Background:** Recent real-world studies have reported significant improvements in the survival of malignant melanoma in the past few years, mainly as a result of modern therapies. However, long-term survival data from Central Eastern European countries such as Hungary are currently lacking.

**Methods:** This nationwide, retrospective study examined melanoma survival in Hungary between 2011–2019 using the databases of the National Health Insurance Fund (NHIF) and Central Statistical Office (CSO) of Hungary. Crude overall survival and age-standardized 5-year net survival as well as the association between age, sex, and survival were calculated.

**Results:** Between 2011 and 2019, 22,948 newly diagnosed malignant melanoma cases were recorded in the NHIF database (47.89% male, mean age: 60.75 years (SD: ±16.39)). 5-year overall survival was 75.40% (women: 80.78%; men: 69.52%). Patients diagnosed between 2017–2019 had a 20% lower risk of mortality compared to patients diagnosed between 2011–2012 (HR 0.80, 95% CI 0.73–0.89; p<0.0001). Age-standardized 5-year net survival rates in 2011–2014 and 2015–2019 were 90.6% and 95.8%, respectively (women: 93.1% and 98.4%, men: 87.8% and 92.7%, respectively). The highest age-standardized 5-year net survival rates were found in the 0–39 age cohort (94.6% in the 2015-2019 period).

**Conclusion:** Hungary has similar melanoma survival rates to Western European countries. Based on net survival, the risk of dying of melanoma within 5 years was cut by more than half (55%) during the study period, which coincides with the successful implementation of awareness campaigns and the wide availability of modern therapies.

## INTRODUCTION

Melanoma has relatively low incidence compared to other non-melanoma cancers of the skin; however, it is responsible for the majority of skin cancer-related mortality (1,2,3,4). Although the number of new cases has been rising in fair-skinned populations and represents significant disease burden (5), some recent publications have indicated a change in trends in mortality over the last decade (6,7). Multiple prognostic factors have been associated with survival in melanoma including age, sex, anatomic site, ulceration, or stage (8,9). Female sex is a positive prognostic factor in melanoma, which is demonstrated by a number of studies showing better survival among women compared to men (10,11). Furthermore, other pathological characteristics such as melanoma thickness (Breslow thickness) are also strongly related to the survival, with thinner lesions having better survival rates than thicker lesions (5).

The incidence and mortality of melanoma varies across European countries. 2018 GLOBOCAN data show wide variations in melanoma incidence and mortality across Europe, with lower incidence and similar mortality rates in Central Eastern European (CEE) countries compared to Western Europe (12). However, apart from differences in UV exposure and preventive activities, the observed lower incidence in CEE countries may also be attributed to suboptimal cancer registration and the lack of centralized cancer registries in some countries (13). Over the past 10 years, survival in advanced melanoma has significantly improved, mainly as a result of modern therapies including immunotherapy and targeted agents (14,15,16,17). The global CONCORD-3 cancer survival surveillance programme compared 5-year net survival rates of melanoma between countries based on registry data. During 2010–2014, 5-year net survival varied between 70%–85% in Central Eastern European countries (CEEC), with modest improvements compared to CONCORD-2 results from 2005–2009 (65%-85%) (18). However, the CONCORD-3 database contains no data from Hungarian registries.

Therefore, our nationwide, retrospective study aimed to investigate 5-year net survival of the total Hungarian melanoma population between 2011–2019, compare it to international results, and evaluate changes in survival during this period. The impact of age and sex on survival as well as on the change of survival rates was also examined.

## MATERIALS AND METHODS

### Data sources

Our nationwide, retrospective study used data from the Hungarian National Health Insurance Fund (NHIF) and the Central Statistical Office (CSO). The NHIF database covers almost the entire Hungarian population and contains information on prescription claims, in- and outpatient visits and medical procedures, as well as medical information using ICD-10 codes. The CSO database provides data on age- and sex-specific mortality from all Hungarian citizens annually. The study was approved by the National Ethical Committee (IV2581-2/2020/EKU). Our study included melanoma patients (ICD-43) diagnosed between January 1, 2011 and December 31, 2019, who were ≥20 years old at the time of diagnosis and had 2 occurrence of C43 ICD-10 codes. A reference period was set from 2009 to 2010 to accurately identify newly diagnosed melanoma patients as it helped to exclude patients with prevalent melanoma at the start of the time window by excluding those with a prior diagnosis code. Patients who died within 60 days of the first C43 code record were also included. Newly diagnosed melanoma patients were followed until December 31, 2019 or until the time of death which was available from the NHIF database without the cause of death. We expressed overall survival (OS) of melanoma population by sex, age and study years based on NHIF database as well as calculated the net survival using NHIF and CSO database inputs using the Pohar Perme estimation.

### Statistical analysis

Data collected were anonymous and non-identifiable. Newly diagnosed melanoma patients are presented as crude incidence numbers on a yearly basis (n). Mean ages were determined for the total time period and for the different diagnostic time intervals (2011–2012, 2013–2014, 2015– 2016, 2017–2019). Crude numbers and mean age are shown for both sexes separately and together. 5-year net survival rates were used for international comparison with CONCORD-3 study results. Net survival corresponds to the cumulative probability of surviving up to a given time since diagnosis (e.g., 5 years) after correcting for other causes of death (as background mortality). Net survival was calculated using the Pohar Perme estimation reported in the CONCORD-3 study, which takes unbiased account of the higher competing risks of death in the elderly (18). We excluded those from the analysis who exceed the age of 95, as Pohar Perme estimation could not calculate 5 year survival within this population. Overall survival is presented using Kaplan-Meier curves. The association between sex, age, and survival was estimated by Cox regression models. We used Cox regression to calculate the age-adjusted hazard ratio (HR) of death of patients diagnosed between 2011–2014 and 2015–2019. All calculations were performed using R version 3.6.1 (05/07/2019) with package boot version 1.3-20.

## RESULTS

### Study population and patient characteristics

In total, 22,948 newly diagnosed malignant melanoma cases were recorded in the NHIF database between 2011 and 2019 (2011–2012: n=4,786; 2013–2014: n=5,060; 2015–2016: n=5,487; 2017–2019: n=7,615) (**Table 1**). In total, 47.89% of patients were male (range: 47.60%–48.13%). The mean age at the time of diagnosis was 60.75 (SD: ±16.39), varying between 63.09 (SD: ±15.20) and 58.01 (SD: ±17.38) years in the male and female population during the investigation period. The highest proportion of cases were found in the 60–69 age group (n=5,445; 23.73% of total), followed by the 70–79 age group (n=5,225; 22.77% of total).

**Table 1.**
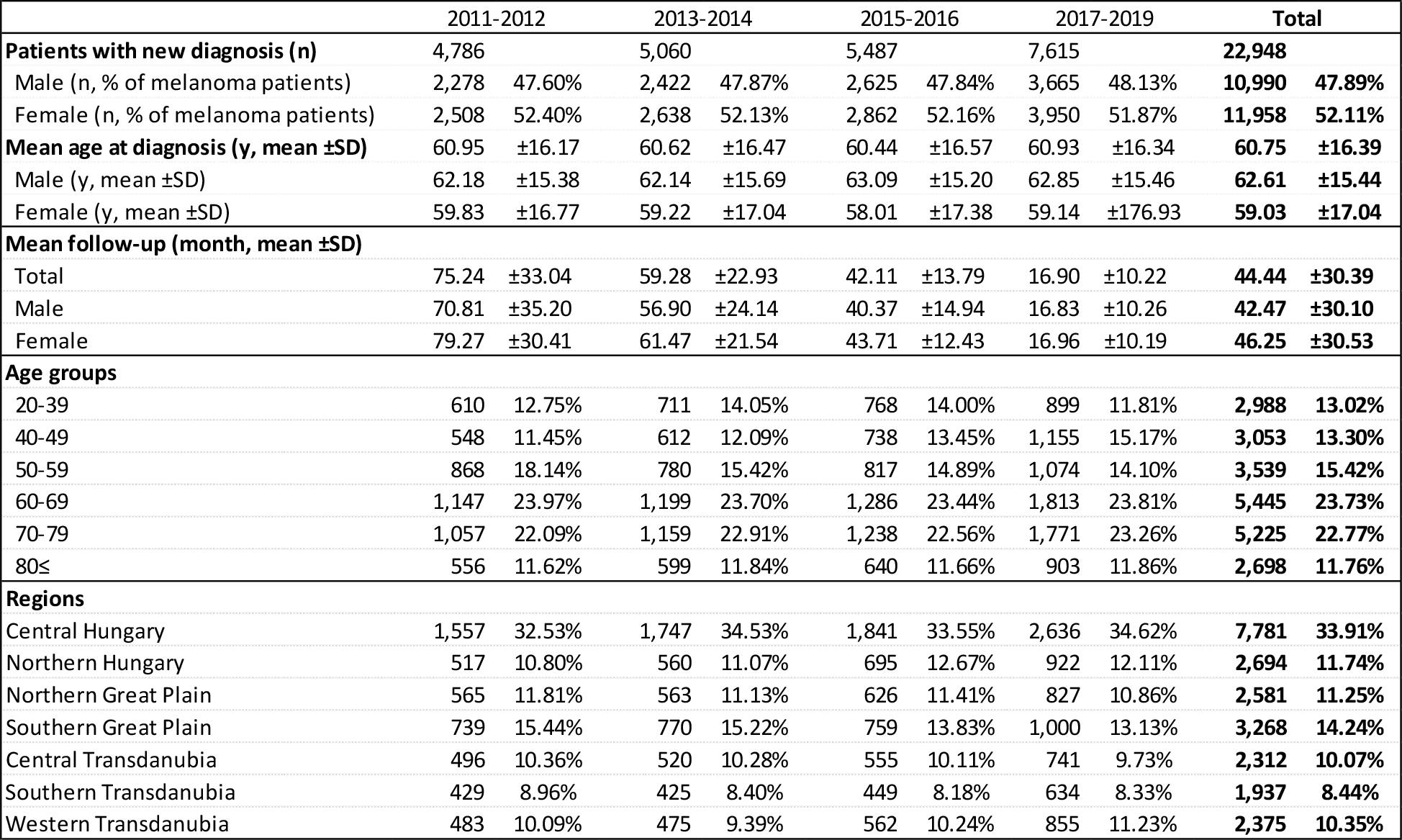
Patient characteristics according to study periods. SD: standard deviation

|  | 2011-2012 |  | 2013-2014 |  | 2015-2016 |  | 2017-2019 |  | Total |  |
| --- | --- | --- | --- | --- | --- | --- | --- | --- | --- | --- |
| **Patients with new diagnosis (n)** | 4,786 |  | 5,060 |  | 5,487 |  | 7,615 |  | **22,948** |  |
| Male (n, % of melanoma patients) | 2,278 | 47.60% | 2,422 | 47.87% | 2,625 | 47.84% | 3,665 | 48.13% | **10,990** | **47.89%** |
| Female (n, % of melanoma patients) | 2,508 | 52.40% | 2,638 | 52.13% | 2,862 | 52.16% | 3,950 | 51.87% | **11,958** | **52.11%** |
| **Mean age at diagnosis (y, mean ±SD)** | 60.95 | $\pm 16.17$ | 60.62 | $\pm 16.47$ | 60.44 | $\pm 16.57$ | 60.93 | $\pm 16.34$ | **60.75** | **$\pm 16.39$** |
| Male (y, mean ±SD) | 62.18 | $\pm 15.38$ | 62.14 | $\pm 15.69$ | 63.09 | $\pm 15.20$ | 62.85 | $\pm 15.46$ | **62.61** | **$\pm 15.44$** |
| Female (y, mean ±SD) | 59.83 | $\pm 16.77$ | 59.22 | $\pm 17.04$ | 58.01 | $\pm 17.38$ | 59.14 | $\pm 176.93$ | **59.03** | **$\pm 17.04$** |
| **Mean follow-up (month, mean ±SD)** |  |  |  |  |  |  |  |  |  |  |
| Total | 75.24 | $\pm 33.04$ | 59.28 | $\pm 22.93$ | 42.11 | $\pm 13.79$ | 16.90 | $\pm 10.22$ | **44.44** | **$\pm 30.39$** |
| Male | 70.81 | $\pm 35.20$ | 56.90 | $\pm 24.14$ | 40.37 | $\pm 14.94$ | 16.83 | $\pm 10.26$ | **42.47** | **$\pm 30.10$** |
| Female | 79.27 | $\pm 30.41$ | 61.47 | $\pm 21.54$ | 43.71 | $\pm 12.43$ | 16.96 | $\pm 10.19$ | **46.25** | **$\pm 30.53$** |
| **Age groups** |  |  |  |  |  |  |  |  |  |  |
| 20-39 | 610 | 12.75% | 711 | 14.05% | 768 | 14.00% | 899 | 11.81% | **2,988** | **13.02%** |
| 40-49 | 548 | 11.45% | 612 | 12.09% | 738 | 13.45% | 1,155 | 15.17% | **3,053** | **13.30%** |
| 50-59 | 868 | 18.14% | 780 | 15.42% | 817 | 14.89% | 1,074 | 14.10% | **3,539** | **15.42%** |
| 60-69 | 1,147 | 23.97% | 1,199 | 23.70% | 1,286 | 23.44% | 1,813 | 23.81% | **5,445** | **23.73%** |
| 70-79 | 1,057 | 22.09% | 1,159 | 22.91% | 1,238 | 22.56% | 1,771 | 23.26% | **5,225** | **22.77%** |
| 80≤ | 556 | 11.62% | 599 | 11.84% | 640 | 11.66% | 903 | 11.86% | **2,698** | **11.76%** |
| **Regions** |  |  |  |  |  |  |  |  |  |  |
| Central Hungary | 1,557 | 32.53% | 1,747 | 34.53% | 1,841 | 33.55% | 2,636 | 34.62% | **7,781** | **33.91%** |
| Northern Hungary | 517 | 10.80% | 560 | 11.07% | 695 | 12.67% | 922 | 12.11% | **2,694** | **11.74%** |
| Northern Great Plain | 565 | 11.81% | 563 | 11.13% | 626 | 11.41% | 827 | 10.86% | **2,581** | **11.25%** |
| Southern Great Plain | 739 | 15.44% | 770 | 15.22% | 759 | 13.83% | 1,000 | 13.13% | **3,268** | **14.24%** |
| Central Transdanubia | 496 | 10.36% | 520 | 10.28% | 555 | 10.11% | 741 | 9.73% | **2,312** | **10.07%** |
| Southern Transdanubia | 429 | 8.96% | 425 | 8.40% | 449 | 8.18% | 634 | 8.33% | **1,937** | **8.44%** |
| Western Transdanubia | 483 | 10.09% | 475 | 9.39% | 562 | 10.24% | 855 | 11.23% | **2,375** | **10.35%** |

Mean follow-up time for those diagnosed between 2011 and 2012 was 75.24 (SD: ±33.04) months, while for those within the 2017-19 period, this value was 16.90 (SD:±10.22) months. Incidence of melanoma was the highest among patients living in Central Hungary Region (33.91% overall) and lowest in Southern Transdanubia Region (8.44%).

### Long-term crude overall survival

At the end of the first year, 92.3% of the total melanoma population was alive (n=22,948). OS rates at 2 years and 5 years were 86.54%, and 75.40%, respectively (**Figure 1**). OS was higher among female patients than male patients throughout the whole study period. 1-year, 2-year, and 5-year OS rates were 93.88%, 89.49%, and 80.78% among female patients, and 90.58%, 83.33%, 69.52% among male patients, respectively. Detailed survival rates of different age cohorts and main Hungarian regions are presented in **Supplementary Table 1**.

**Figure 1.**
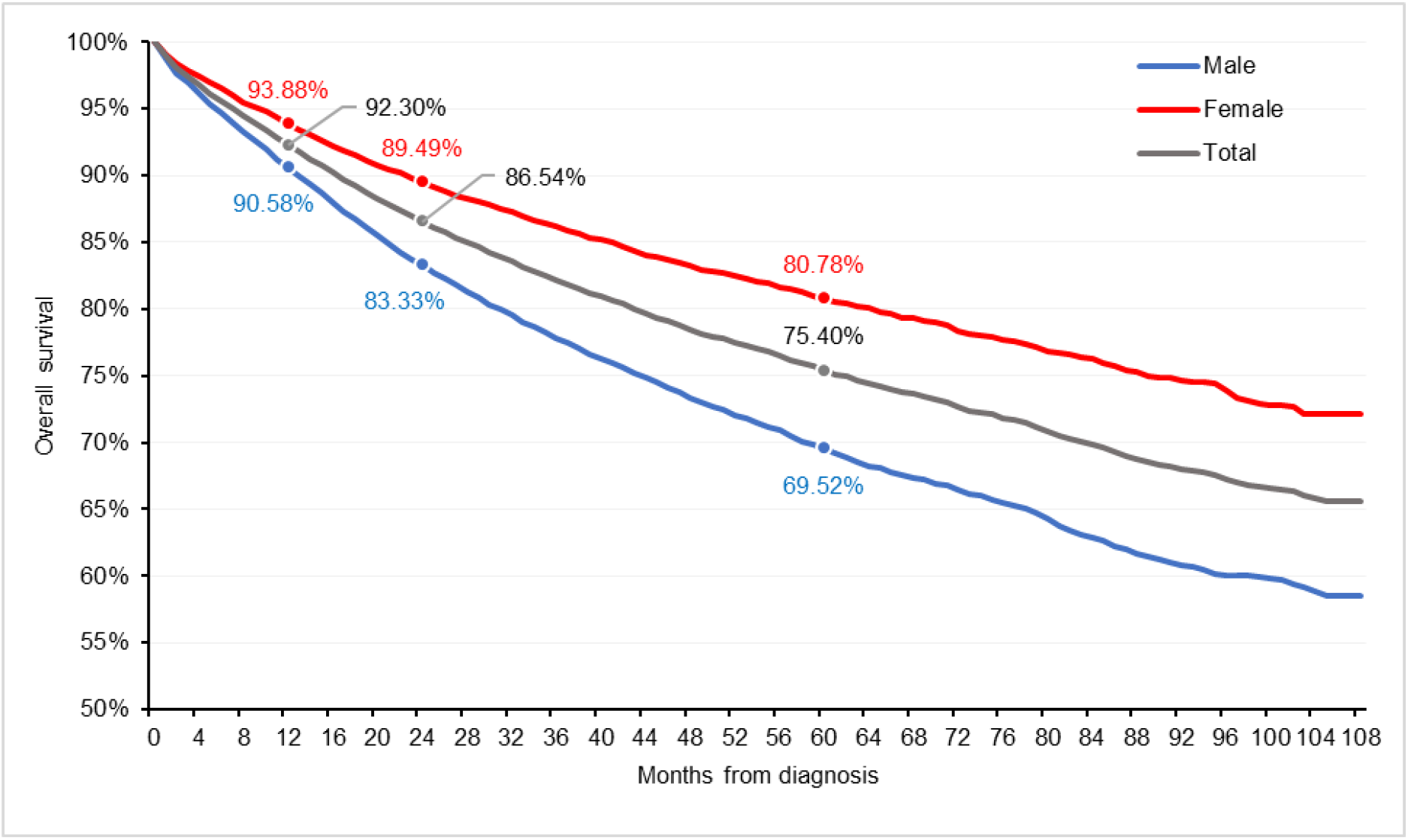
Estimated overall survival of Hungarian malignant melanoma patients between 2011 and 2019

### Change in crude overall survival between 2011–2012 and 2017–2019

**Figure 2** shows overall survival among patients diagnosed between 2011–2012 and 2017–2019. In the 2011–2012 period, newly diagnosed melanoma patients had 1-year, 2-year, and 3-year OS rates of 90.85%, 84.66%, and 80.78%, respectively. Corresponding values in the 2017–2019 period were 92.90%, 87.28%, and 83.52%, respectively.

**Figure 2.**
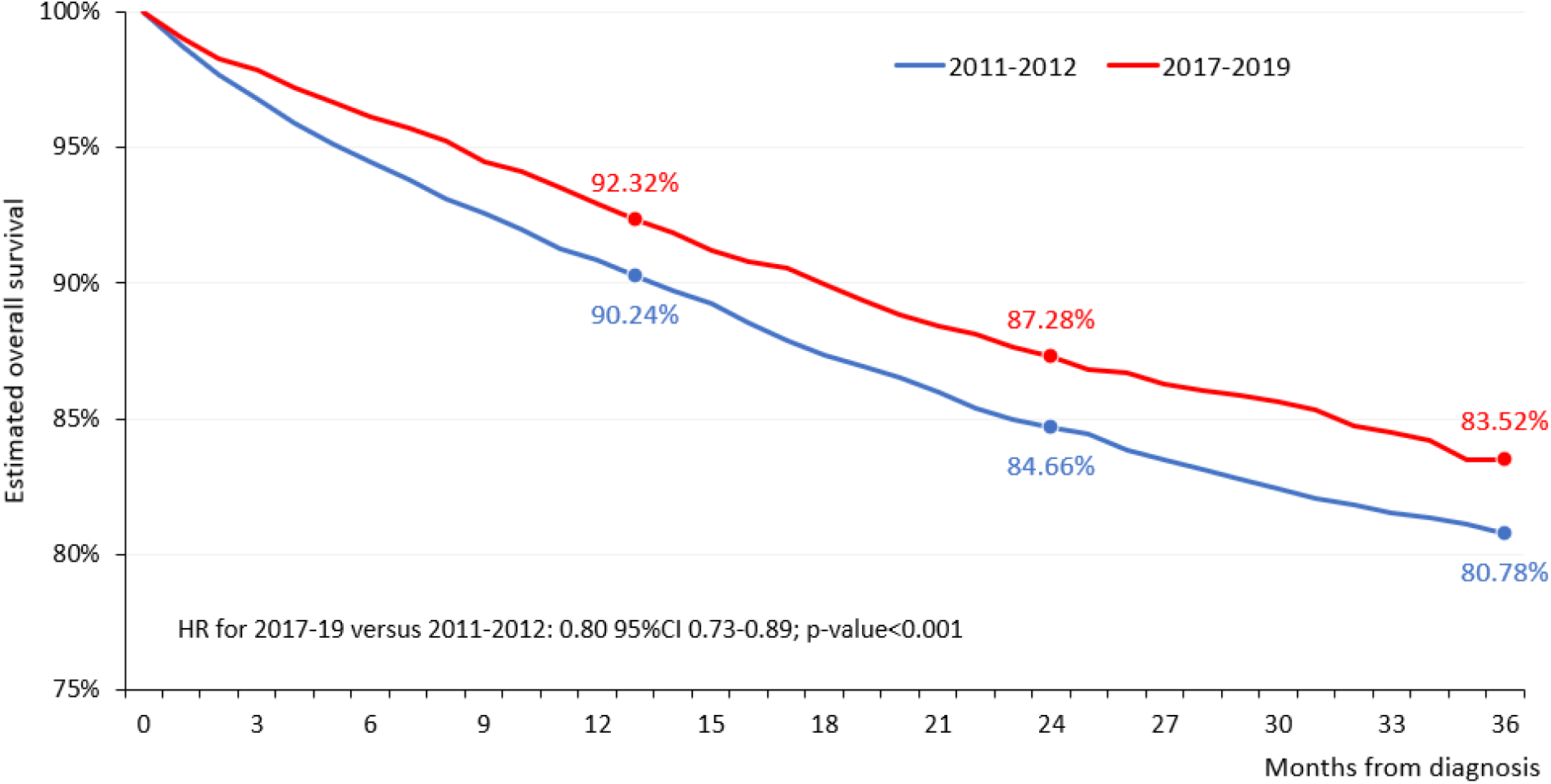
Overall survival of melanoma patients diagnosed between 2011-12 and 2017-2019

Melanoma patients diagnosed between 2017–2019 had a 20% lower risk of mortality compared to patients diagnosed between 2011–2012 (HR 0.80, 95% CI 0.73–0.89; p<0.0001). Male patients had a larger decrease in mortality (HR 0.78, 95% CI 0.69–0.89; p<0.001) compared to female patients (HR 0.84, 95% CI 0.72-0.98; p=0.024) **(Figure 3)**. The largest reduction in mortality was seen in the 50–59 age cohort (HR 0.43, 95% CI 0.31–0.61; <0.0001); the 60–69 and 85+ age cohorts had non-significant decreases in mortality. The largest mortality reduction was found among patients living in the Central Transdanubian region (HR 0.65, 95% CI 0.43-0.98, p=0.04), and non-significant decreases were seen in Northern Hungary, the Southern Great Plain, and Southern Transdanubia.

**Figure 3.**
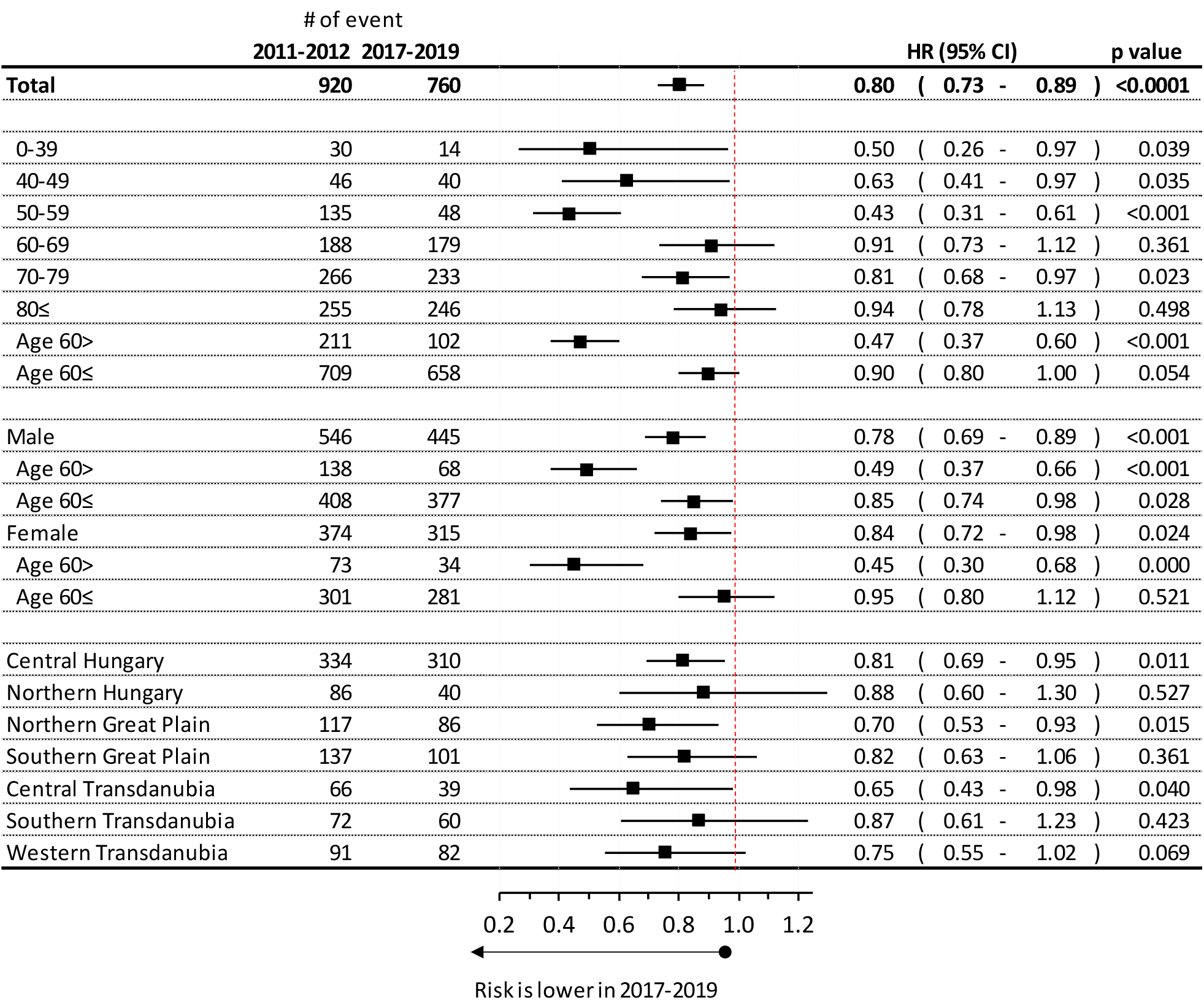
Hazard ratio of overall survival of melanoma patients by age and sex diagnosed between 2011-12 and 2017-2019

### Age-standardized 5-year net survival of Hungarian melanoma patients

Age-standardized 5-year net survival rates in 2011–2014 and 2015–2019 were 90.59% and 95.78%, respectively. Corresponding values were 93.13% and 98.36% among female and 87.78% and 92.67% among male patients, respectively (**Figure 4**). The increase in net survival between the 2015-2019 and 2011-2014 periods was significant in the case of the total and female populations (p-value<0.001at both cases), however did not reach statistical significance in the male melanoma population (p=0.095).

**Figure 4.**
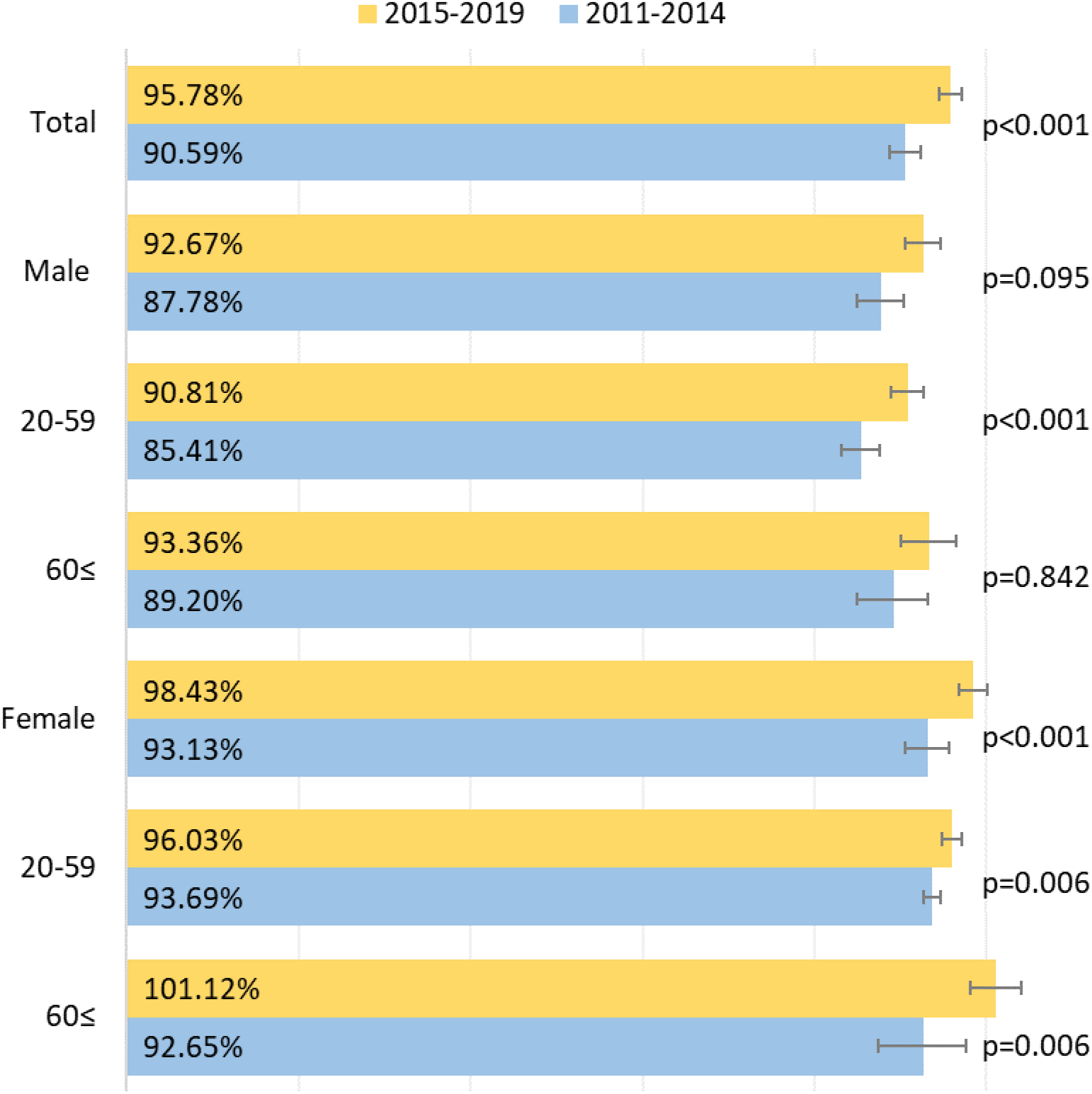
Age-standardized 5-year net survival of Hungarian melanoma patients overall and by sex diagnosed between 2011–2014 and 2015–2019

Age-standardized 5-year net survival of age cohorts and main Hungarian regions are detailed in **Supplementary Table 2**.

## DISCUSSION

Our nationwide, retrospective study was the first to examine long-term survival of melanoma in Hungary. Results show that melanoma net survival in Hungary was similar to that of Western European countries during the early 2010’s. We demonstrated a significant mortality reduction of melanoma patients between the 2011–2012 and 2017–2019 periods. Furthermore, we also found that female sex and younger age were associated with better survival. Age-standardized 5-year net survival of melanoma also improved considerably during the study period in total and for females.

Improvements in melanoma survival have been reported by a number of recent real-world studies (14,19,20,21,22), the reasons for which are multifactorial. First, the improvement of diagnostic modalities has allowed for an earlier detection of the disease and thus the timely initiation of life-prolonging therapy. Furthermore, the widespread implementation of melanoma awareness campaigns and screening programs worldwide has also contributed to earlier detection and better prognosis. Australia, one of the most affected countries, has conducted a number of successful educational and awareness campaigns starting from the 1980s (23). In the randomized Healthy Text study, theory-based text messages about skin cancer prevention and skin self-examination resulted in significant improvements in sun-protection and skin self-examination behaviors among people aged 18–42 years (24). The SunSmart skin cancer prevention program was initiated in 1988 with a view to raise public awareness, promote preventive behaviors and environmental change for skin cancer prevention. SunSmart resulted in a rapid increase in the use of sun protection and contributed to a reduction in melanoma in younger age groups (25,). In Europe, the Euromelanoma Skin Cancer Prevention Campaign was founded in 1999 by six Belgian dermatologists and is now active in 33 European countries with public awareness campaigns and melanoma screening days, scientific publications and special events (www.euromelanoma.org). The Euromelanoma initiative has raised greater awareness of melanoma and increased the number of screenings and the detection of clinically suspected lesions (26,27). Increasing community awareness of melanoma and its risk factors was associated with a progressive decreasing incidence of thicker melanoma and for a long-term decrease in mortality according to the experience of some countries such as Germany, Australia and the United States (22).

Accordingly, the majority of recent real-world studies have directly linked improvements in melanoma survival to the introduction of modern therapies (14,16,2,21). We found better overall survival rates among female vs. male patients in the Hungarian melanoma population during the whole study period, which is also in line with previous observations (28,29).

To our knowledge, our study is the first to report 5-year net survival rates of melanoma in Hungary based on the database of the CSO. Age-standardized 5-year net survival of melanoma increased from 90.6% in 2011–2014 to 95.8% in 2015–2019, with higher survival rates observed in women and in younger age cohorts during the whole study period. To compare our results with findings from other countries, we examined data from the worldwide CONCORD cancer survival surveillance program for the period of 2011–2014 (18). Age-standardized 5-year net survival of melanoma in Hungary was comparable to that of Western and Northern European countries including Denmark, Sweden, the United Kingdom, Belgium, France, Germany, the Netherlands, and Switzerland where 5-year survival rates also exceeded 90% in this period. This is in line with our previous findings showing that melanoma mortality in Hungary is close to the European average (6) and reflects the effectiveness of the successful local implementation of the Euromelanoma program in 2009 (30). In the interest of high-quality comprehensive cancer control, Hungarian healthcare organisations implemented recommendations for high-quality patient care in melanoma management according to progressivity (31) and nominated 7 clinics as “Melanoma Centres” to promote clinical experience with modern immune and targeted therapies (32). Furthermore, almost the entirety of the Hungarian population is covered by social insurance with full access to reimbursed therapies and has equity in patient care.

The favourable changes in mortality may be attributed to several factors including the relatively high and early access to innovative therapies and high health expenditure per capita compared to neighboring European countries (33). In addition, earlier detection may also contribute to improvements in prognosis, which is supported by a study reporting a significant decrease in mean Breslow thickness from 2.2 mm in 1998 to 1.6 mm in 2008 (34). Finally, the wide availability of modern oncologic diagnostic and treatment modalities in Hungary is likely a key driver of favorable melanoma outcomes. The introduction of modern therapies has dramatically changed the landscape for melanoma (35,36). Ipilimumab was the first immunotherapy agent approved for the treatment of irresectable, metastatic melanoma by the European Medicines Agency (EMA) in 2012 and followed by the first targeted therapy in 2014. Immunotherapy has revolutionized the treatment of cancers. Pembrolizumab, nivolumab and nivolumab/ipilimumab combination therapy gained approvals in melanoma and in several tumor types over time (37,38). These innovative drugs became available in Hungary shortly after their approval in the European Union, and the period of their introduction coincides with a period of similar survival improvements in Hungary compared to other European countries.

We found higher 5-year net survival rates among female melanoma patients compared to men throughout the whole study period. Our findings are consistent with those reported by the Cancer Research UK database which reported 5-year net survival rates of 92.4% in women and 87.6% in men in the 2010–2011 period (39). The survival advantage of women in melanoma is well-established (10,40,41). A recent large analysis from the U.S. Surveillance, Epidemiology, and End Results (SEER) cancer database showed significantly better 3- and 5-year cancer-specific melanoma survival rates among women compared to men (40), and Behbanani *et al*. also reported better 5-year overall survival for female melanoma patients based on the U.S. National Cancer Registry (10). The positive prognostic significance of female sex may be explained by various factors. Several reports suggest that gender disparities in melanoma survival may partly be due to gender-related differences in skin cancer risk awareness, protection against UV exposure, and self-examination (42,43,44). Furthermore, women have been shown to have a slower rate of growth of the primary tumor and a lower risk of metastasis (45). Recent reports suggest that apart from differences in gender-related behavior, genetic and epigenetic aspects as well as environmental and biological factors may also play an important part in the positive prognostic significance of female sex in melanoma survival (46,47). In our earlier report, we demonstrated the Hungarian mortality-to-incidence ratios showed a decrease over the past decade in both sexes and reached the level of Western and Northern European countries, although males had higher MIRs in all study years in comparison with women (48).

Our study corresponds well with the growing body of evidence showing gender-related differences in melanoma survival in the era of modern therapies and warrants further investigations into potentially modifiable risk factors responsible for poorer prognosis among men compared to women.

In our study, younger age groups had better 5-year net survival rates than older patients. Specifically, the highest survival rate was seen for patients aged 0–39 years (95%), while the lowest rate was found for the 70–79 age cohort (87.3%). The better prognosis of melanoma among younger ages is well-documented (49), and our findings are also in line with data from the Cancer Research UK database showing a continuous decrease in 5-year net overall survival with increasing age (50). While better survival rates in younger ages may partly be explained by the generally better health status of these age groups, several reports suggest that other factors also play a part in the survival gap. First, older patients present with significantly thicker melanomas, a higher mitotic rate and increased incidence of ulceration compared to young patients (51,52,53). Although a declining trend in Breslow thickness has been reported recently, the decline seems to be less prominent in older patients compared to young patients, reflecting poorer prognosis for the elderly (52). Besides melanoma thickness, older patients are more often diagnosed with head and neck melanomas and nodular lesions which develop very quickly and are associated with poor prognosis (54). Elderly patients may have gained less advantage from public health campaigns and thus have lower risk awareness and thus later (self-)detection (51), and suboptimal staging and/or treatment may also contribute to poorer prognosis (54). Furthermore, they are often not considered candidates for surgical treatment, resulting in a lower excision rate and longer time to definitive excision of primary melanoma (54,55). Besides, aging of immune system could contribute to the higher incidence of malignancies, more aggressive form of certain cancer types and worse response for given therapies, which could result in worse outcomes (56). In summary, there is still room for improvement in melanoma management among the elderly to ensure that they benefit more from newly available treatment options.

The main strengths and limitations of our analysis both lie in the nature of the NHIF database, which allowed for the inclusion of all Hungarian melanoma patients diagnosed between 2011 and 2019, however does not contain any information on patient clinical characteristics, vital signs, laboratory findings, or baseline prognostic features.

## CONCLUSION

This study was the first to examine long-term overall survival and 5-year net survival of melanoma in Hungary. The results show that melanoma survival in Hungary is similar to that of Western European countries. We found better survival rates in women and younger age groups, and based on net survival, the risk of dying of melanoma within 5 years was cut by more than half (55%) during the study period, which coincides with the successful implementation of awareness campaigns and the wide availability of modern therapies.

## Data Availability

The raw data supporting the conclusions of this article will be made available by the authors, without undue reservation.

## ACKNOWLEDGEMENTS

We would like to thank the NHIF and CSO as well as the National Cancer Registry for providing comprehensive datasets for our analysis. We would also like to thank Zsófia Barcza of Syntesia Medical Communications for medical writing support.

## FUNDING

The authors declare that this study received funding from MSD Pharma Hungary. The funder had the following involvement with the study: in study design, data collection and analysis, decision to publish, and preparation of the manuscript.

## CONFLICT OF INTEREST STATEMENT

Authors ZK, ZP, MV, AB and KK were employed by the company MSD Pharma Hungary. MV, BN and ZV are employees of Semmelweis University. Semmelweis University received a grant from MSD Pharma Hungary to contribute to this research. GyR and IF are employees of RxTarget Ltd. and ZsB is employed of Syntesia Ltd. where their contribution to this project was financially compensated. The remaining authors declare that the research was conducted in the absence of any commercial or financial relationships that could be construed as a potential conflict of interest.

## AVAILABILITY OF DATA AND MATERIALS

Data sharing is available in supplementary tables and figures, and the authors are happy to share all available data from the study.

## Supplementary Table

**Supplementary Table 1.**
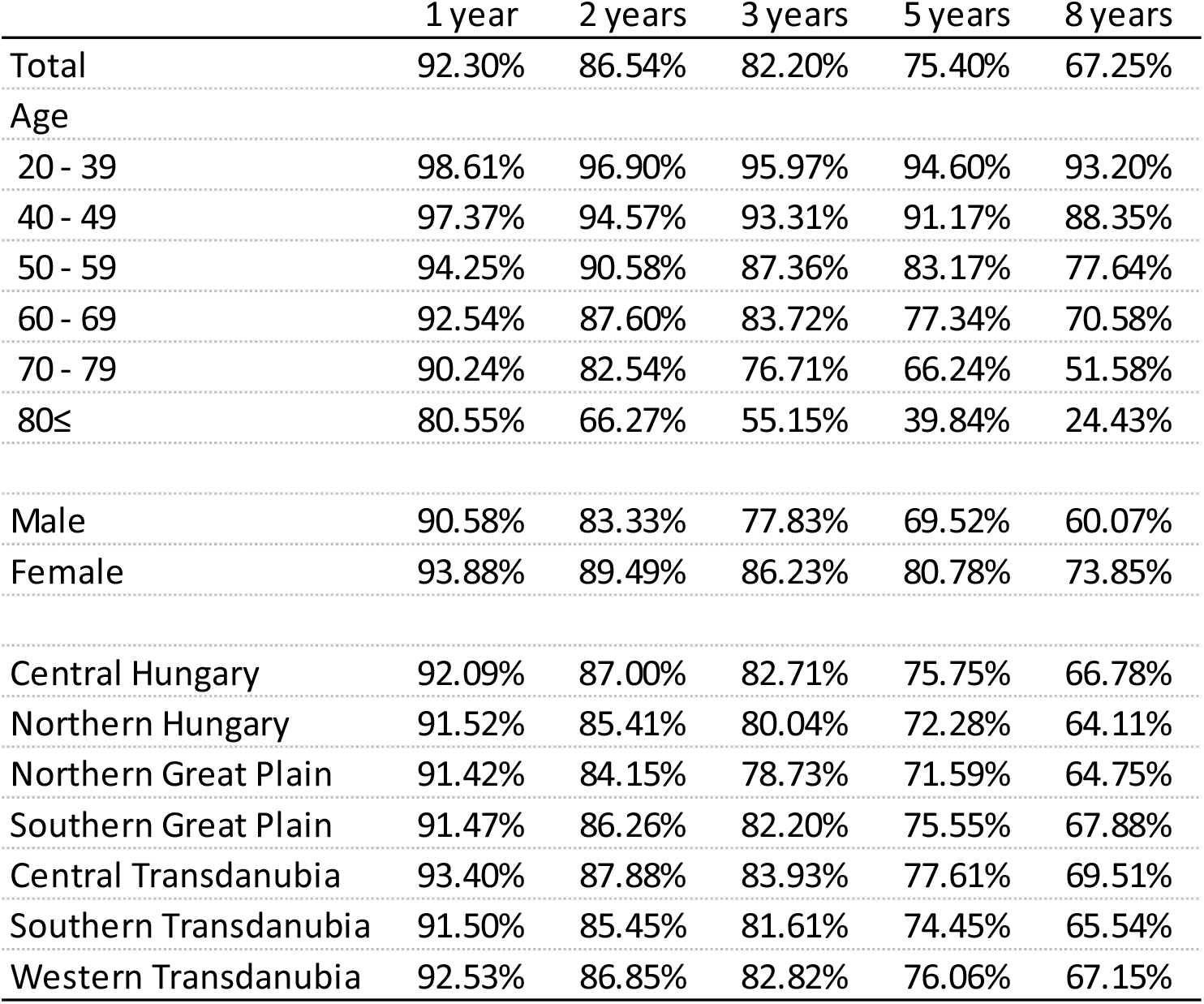
Estimated overall survival of Hungarian malignant melanoma patients between 2011 and 2019 by age, sex and main Hungarian regions

**Supplementary Table 2.**
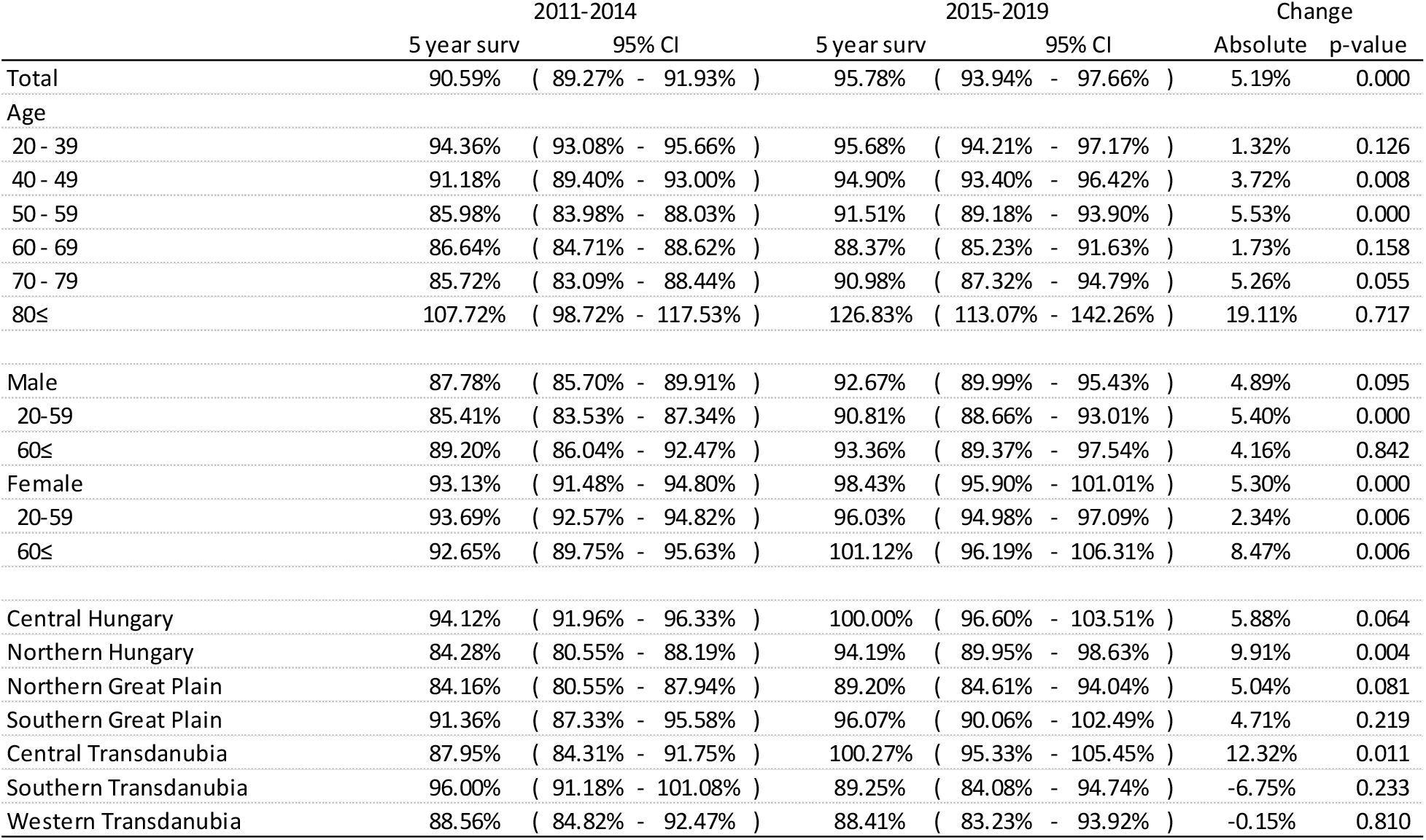
Age-standardized 5-year net survival of Hungarian melanoma patients overall, by sex, age cohorts and main regions diagnosed between 2011–2014 and 2015–2019

## Notes

### Competing Interest Statement

The authors declare a potential conflict of interest as listed below: Authors ZK, ZP, MV, AB and KK were employed by the company MSD Pharma Hungary. MV, BN and ZV are employees of Semmelweis
University. Semmelweis University received a grant from MSD Pharma Hungary to contribute to this research. GyR and IF are employees of RxTarget Ltd. and ZsB is employed of Syntesia Ltd. where their contribution to this project was financially
compensated. The remaining authors declare that the research was conducted in the absence of any commercial or financial relationships that could be construed as a potential conflict of interest.

### Author Declarations

The Medical Research Council in Hungary (Mailing address: 7-8 Szechenyi Istvan ter, Budapest, H-1051, Hungary) gave ethical approval for this work (IV2581-2/2020/EKU).

