## Supplementary material for "SIGNIFICANT IMPROVEMENT IN MELANOMA SURVIVAL OVER THE LAST DECADE: A HUNGARIAN NATIONWIDE STUDY BETWEEN 2011–2019": Table 1

Table 1 Patient characteristics according to study periods. SD: standard deviation

|  | 2011-2012 | | 2013-2014 | | 2015-2016 | | 2017-2019 | | **Total** | |
| --- | --- | --- | --- | --- | --- | --- | --- | --- | --- | --- |
| **Patients with new diagnosis (n)** | 4,786 |  | 5,060 |  | 5,487 |  | 7,615 |  | **22,948** |  |
| Male (n, % of LC patients) | 2,278 | 47.60% | 2,422 | 47.87% | 2,625 | 47.84% | 3,665 | 48.13% | **10,990** | **47.89%** |
| Female (n, % of LC patients) | 2,508 | 52.40% | 2,638 | 52.13% | 2,862 | 52.16% | 3,950 | 51.87% | **11,958** | **52.11%** |
| **Mean age at diagnosis (y, mean ±SD)** | 60.95 | ±16.17 | 60.62 | ±16.47 | 60.44 | ±16.57 | 60.93 | ±16.34 | **60.75** | **±16.39** |
| Male (y, mean ±SD) | 62.18 | ±15.38 | 62.14 | ±15.69 | 63.09 | ±15.20 | 62.85 | ±15.46 | **62.61** | **±15.44** |
| Female (y, mean ±SD) | 59.83 | ±16.77 | 59.22 | ±17.04 | 58.01 | ±17.38 | 59.14 | ±176.93 | **59.03** | **±17.04** |
| **Mean follow-up (month, mean ±SD)** |  |  |  |  |  |  |  |  |  |  |
| Total | 75.24 | ±33.04 | 59.28 | ±22.93 | 42.11 | ±13.79 | 16.90 | ±10.22 | **44.44** | **±30.39** |
| Male | 70.81 | ±35.20 | 56.90 | ±24.14 | 40.37 | ±14.94 | 16.83 | ±10.26 | **42.47** | **±30.10** |
| Female | 79.27 | ±30.41 | 61.47 | ±21.54 | 43.71 | ±12.43 | 16.96 | ±10.19 | **46.25** | **±30.53** |
| **Age groups** |  |  |  |  |  |  |  |  |  |  |
| 20-39 | 610 | 12.75% | 711 | 14.05% | 768 | 14.00% | 899 | 11.81% | **2,988** | **13.02%** |
| 40-49 | 548 | 11.45% | 612 | 12.09% | 738 | 13.45% | 1,155 | 15.17% | **3,053** | **13.30%** |
| 50-59 | 868 | 18.14% | 780 | 15.42% | 817 | 14.89% | 1,074 | 14.10% | **3,539** | **15.42%** |
| 60-69 | 1,147 | 23.97% | 1,199 | 23.70% | 1,286 | 23.44% | 1,813 | 23.81% | **5,445** | **23.73%** |
| 70-79 | 1,057 | 22.09% | 1,159 | 22.91% | 1,238 | 22.56% | 1,771 | 23.26% | **5,225** | **22.77%** |
| 80≤ | 556 | 11.62% | 599 | 11.84% | 640 | 11.66% | 903 | 11.86% | **2,698** | **11.76%** |
| **Regions** |  |  |  |  |  |  |  |  |  |  |
| Central Hungary | 1,557 | 32.53% | 1,747 | 34.53% | 1,841 | 33.55% | 2,636 | 34.62% | **7,781** | **33.91%** |
| Northern Hungary | 517 | 10.80% | 560 | 11.07% | 695 | 12.67% | 922 | 12.11% | **2,694** | **11.74%** |
| Northern Great Plain | 565 | 11.81% | 563 | 11.13% | 626 | 11.41% | 827 | 10.86% | **2,581** | **11.25%** |
| Southern Great Plain | 739 | 15.44% | 770 | 15.22% | 759 | 13.83% | 1,000 | 13.13% | **3,268** | **14.24%** |
| Central Transdanubia | 496 | 10.36% | 520 | 10.28% | 555 | 10.11% | 741 | 9.73% | **2,312** | **10.07%** |
| Southern Transdanubia | 429 | 8.96% | 425 | 8.40% | 449 | 8.18% | 634 | 8.33% | **1,937** | **8.44%** |
| Western Transdanubia | 483 | 10.09% | 475 | 9.39% | 562 | 10.24% | 855 | 11.23% | **2,375** | **10.35%** |
