## Supplementary Table 1 for "SIGNIFICANT IMPROVEMENT IN MELANOMA SURVIVAL OVER THE LAST DECADE: A HUNGARIAN NATIONWIDE STUDY BETWEEN 2011–2019"

Supplementary Table 1 Estimated overall survival of Hungarian malignant melanoma patients between 2011 and 2019 by age, sex and main Hungarian regions

|  | 1 year | 2 years | 3 years | 5 years | 8 years |
| --- | --- | --- | --- | --- | --- |
| Total | 92.30% | 86.54% | 82.20% | 75.40% | 67.25% |
| Age |  |  |  |  |  |
| 20 - 39 | 98.61% | 96.90% | 95.97% | 94.60% | 93.20% |
| 40 - 49 | 97.37% | 94.57% | 93.31% | 91.17% | 88.35% |
| 50 - 59 | 94.25% | 90.58% | 87.36% | 83.17% | 77.64% |
| 60 - 69 | 92.54% | 87.60% | 83.72% | 77.34% | 70.58% |
| 70 - 79 | 90.24% | 82.54% | 76.71% | 66.24% | 51.58% |
| 80≤ | 80.55% | 66.27% | 55.15% | 39.84% | 24.43% |
| Male | 90.58% | 83.33% | 77.83% | 69.52% | 60.07% |
| Female | 93.88% | 89.49% | 86.23% | 80.78% | 73.85% |
| Central Hungary | 92.09% | 87.00% | 82.71% | 75.75% | 66.78% |
| Northern Hungary | 91.52% | 85.41% | 80.04% | 72.28% | 64.11% |
| Northern Great Plain | 91.42% | 84.15% | 78.73% | 71.59% | 64.75% |
| Southern Great Plain | 91.47% | 86.26% | 82.20% | 75.55% | 67.88% |
| Central Transdanubia | 93.40% | 87.88% | 83.93% | 77.61% | 69.51% |
| Southern Transdanubia | 91.50% | 85.45% | 81.61% | 74.45% | 65.54% |
| Western Transdanubia | 92.53% | 86.85% | 82.82% | 76.06% | 67.15% |
