## Supplementary Table 2 for "SIGNIFICANT IMPROVEMENT IN MELANOMA SURVIVAL OVER THE LAST DECADE: A HUNGARIAN NATIONWIDE STUDY BETWEEN 2011–2019"

Supplementary Table 2 Age-standardized 5-year net survival of Hungarian melanoma patients overall, by sex, age cohorts and main regions diagnosed between 2011–2014 and 2015–2019

|  | 2011-2014 | | | | | |  | 2015-2019 | | | | | |  | Change | |
| --- | --- | --- | --- | --- | --- | --- | --- | --- | --- | --- | --- | --- | --- | --- | --- | --- |
|  | 5 year surv |  | 95% CI | | |  |  | 5 year surv |  | 95% CI | | | | | Absolute | p-value |
| Total | 90.59% | ( | 89.27% | - | 91.93% | ) |  | 95.78% | ( | 93.94% | - | 97.66% | ) |  | 5.19% | 0.000 |
| Age |  |  |  |  |  |  |  |  |  |  |  |  |  |  |  |  |
| 20 - 39 | 94.36% | ( | 93.08% | - | 95.66% | ) |  | 95.68% | ( | 94.21% | - | 97.17% | ) |  | 1.32% | 0.126 |
| 40 - 49 | 91.18% | ( | 89.40% | - | 93.00% | ) |  | 94.90% | ( | 93.40% | - | 96.42% | ) |  | 3.72% | 0.008 |
| 50 - 59 | 85.98% | ( | 83.98% | - | 88.03% | ) |  | 91.51% | ( | 89.18% | - | 93.90% | ) |  | 5.53% | 0.000 |
| 60 - 69 | 86.64% | ( | 84.71% | - | 88.62% | ) |  | 88.37% | ( | 85.23% | - | 91.63% | ) |  | 1.73% | 0.158 |
| 70 - 79 | 85.72% | ( | 83.09% | - | 88.44% | ) |  | 90.98% | ( | 87.32% | - | 94.79% | ) |  | 5.26% | 0.055 |
| 80≤ | 107.72% | ( | 98.72% | - | 117.53% | ) |  | 126.83% | ( | 113.07% | - | 142.26% | ) |  | 19.11% | 0.717 |
| Male | 87.78% | ( | 85.70% | - | 89.91% | ) |  | 92.67% | ( | 89.99% | - | 95.43% | ) |  | 4.89% | 0.095 |
| 20-59 | 85.41% | ( | 83.53% | - | 87.34% | ) |  | 90.81% | ( | 88.66% | - | 93.01% | ) |  | 5.40% | 0.000 |
| 60≤ | 89.20% | ( | 86.04% | - | 92.47% | ) |  | 93.36% | ( | 89.37% | - | 97.54% | ) |  | 4.16% | 0.842 |
| Female | 93.13% | ( | 91.48% | - | 94.80% | ) |  | 98.43% | ( | 95.90% | - | 101.01% | ) |  | 5.30% | 0.000 |
| 20-59 | 93.69% | ( | 92.57% | - | 94.82% | ) |  | 96.03% | ( | 94.98% | - | 97.09% | ) |  | 2.34% | 0.006 |
| 60≤ | 92.65% | ( | 89.75% | - | 95.63% | ) |  | 101.12% | ( | 96.19% | - | 106.31% | ) |  | 8.47% | 0.006 |
| Central Hungary | 94.12% | ( | 91.96% | - | 96.33% | ) |  | 100.00% | ( | 96.60% | - | 103.51% | ) |  | 5.88% | 0.064 |
| Northern Hungary | 84.28% | ( | 80.55% | - | 88.19% | ) |  | 94.19% | ( | 89.95% | - | 98.63% | ) |  | 9.91% | 0.004 |
| Northern Great Plain | 84.16% | ( | 80.55% | - | 87.94% | ) |  | 89.20% | ( | 84.61% | - | 94.04% | ) |  | 5.04% | 0.081 |
| Southern Great Plain | 91.36% | ( | 87.33% | - | 95.58% | ) |  | 96.07% | ( | 90.06% | - | 102.49% | ) |  | 4.71% | 0.219 |
| Central Transdanubia | 87.95% | ( | 84.31% | - | 91.75% | ) |  | 100.27% | ( | 95.33% | - | 105.45% | ) |  | 12.32% | 0.011 |
| Southern Transdanubia | 96.00% | ( | 91.18% | - | 101.08% | ) |  | 89.25% | ( | 84.08% | - | 94.74% | ) |  | -6.75% | 0.233 |
| Western Transdanubia | 88.56% | ( | 84.82% | - | 92.47% | ) |  | 88.41% | ( | 83.23% | - | 93.92% | ) |  | -0.15% | 0.810 |
